# A One Health Investigation into H5N1 Avian Influenza Virus Epizootics on Two Dairy Farms

**DOI:** 10.1101/2024.07.27.24310982

**Authors:** Ismaila Shittu, Diego Silva, Judith U. Oguzie, Lyudmyla V. Marushchak, Gene G. Olinger, John A. Lednicky, Claudia M. Trujillo-Vargas, Nicholas E. Schneider, Haiping Hao, Gregory C. Gray

## Abstract

**Background:** In early April 2024 we studied two Texas dairy farms which had suffered incursions of H5N1 highly pathogenic avian influenza virus (HPAIV) the previous month.

**Methods:** We employed molecular assays, cell and egg culture, Sanger and next generation sequencing to isolate and characterize viruses from multiple farm specimens (cow nasal swab, milk specimens, fecal slurry, and a dead bird).

**Results:** We detected H5N1 HPAIV in 64% (9/14) of milk specimens, 2.6% (1/39) of cattle nasal swab specimens, and none of 17 cattle worker nasopharyngeal swab specimens. We cultured and characterized virus from eight H5N1-positive specimens. Sanger and next-generation sequencing revealed the viruses were closely related into other recent Texas epizootic H5N1 strains of clade 2.3.4.4b. Our isolates had multiple mutations associated with increased spillover potential. Surprisingly, we detected SARS-CoV-2 in a nasal swab from a sick cow. Additionally, 14.3% (2/14) of the farm workers who donated sera were recently symptomatic and had elevated neutralizing antibodies against a related H5N1 strain.

**Conclusions:** While our sampling was limited, these data offer additional insight into the large H5N1 HPAIV epizootic which thus far has impacted at least 96 cattle farms in twelve US states. Due to fears that research might damage dairy businesses, studies like this one have been few. We need to find ways to work with dairy farms in collecting more comprehensive epidemiological data that are necessary for the design of future interventions against H5N1 HPAIV on cattle farms.

---

Highly pathogenic avian influenza A (HPAIV) subtype (H5N1) viruses continue to diversify genetically and have caused millions of wild bird and poultry deaths across multiple continents^1,2^. Most recently, HPAIV H5N1 strains have infected numerous animal species including bears, bobcats, coyotes, foxes, goats, racoons, sea lions, skunks, and most recently, cattle ^3,4^. Particularly prevalent in these spillovers event have been HPAIV H5N1 HA clade 2.3.3.4b^5,6^.

On the 25^th^ of March 2024^7^, the USDA confirmed an outbreak of HPAIV H5N1 in a dairy farm in Texas. As we write this draft (July 25, 2024) the USDA’s farm count is currently at 172 cattle farms^8^ in 13 states. Four dairy farm and nine poultry farm workers are thought to have been recently infected from these viral strains. While influenza A viruses have been previously detected in cattle^9^, this is the first instance of such widespread infections spreading rapidly across multiple different geographical areas. Upon invitation we used a previously approved One Health research protocol to study two Texas dairy farms for novel respiratory viruses.

## Methods

### Study Sites and Sampling Protocol

Knowing that we had a research proposal to study livestock farms for evidence of novel respiratory viruses, we were invited by farm owners to study two dairy farms in Texas as they were recovering from incursions of avian influenza A H5N1 virus in their cattle. The identity and locations of two dairy farms (Farm A and B) are protected through nondisclosure agreements.

Our protocol was approved by the University of Texas Medical Branch’s (UTMB) Institutional Review Board (23-0085). UTMB’s Institutional Animal Care and Use Committee (IACUC) viewed this study as exempt from formal IACUC ethical review. Our One Health study called for studying livestock farms in United States and Mexico for novel respiratory viruses.^10–12^ We planned to prospectively sample sick and healthy farm livestock, the farm environments, and a cohort of healthy animal workers for evidence of novel coronaviruses and other respiratory viruses. At enrollment and every four months for a total of 12 months, we planned to collect questionnaire data about each farm and its animal workers, and an array of samples: 20 nasal swabs from livestock (up to 70% from animals with signs of respiratory illness), NP swabs and sera from up to 10 animal workers, and 4 three-hour bioaerosol samples. In between the four planned farm visits, farm employees would use postage-paid sample kits to collect and ship nasal swabs from livestock with signs of respiratory illness.

We employed classical techniques in field sample preparation and preservation. More detailed information can be found in the appendix.

### Human Samples

Prior to sampling, farm workers of at least 18 years in age were invited to participate in the study through informed consent. Each worker was given a questionnaire to gather demographic and other relevant information related to routine daily activities on the farm. Up to 10 farm workers per farm were asked to permit a serum and a nasopharyngeal (NP) swabs collection.

### Animals Samples

Farm staff identified as many as 14 sick cattle and 6 healthy cattle at each farm and collected a deep nasal swab sample from each animal. Data regarding the cattle were captured on a sampling form. Additionally, at Farm B we were invited to collect milk samples from a group of previously ill cattle, and orotracheal and cloacal swab samples from the dead grackle found that morning in a cattle barn.

### Environmental Samples

We used bioaerosol cyclone samplers to study the farms for evidence of novel viruses. We studied areas such as milking parlors, areas where cows queued up to be milked, and hospital pens. In addition, a manager on Farm B collected a fecal slurry sample and asked us to examine it for novel viruses.

### Laboratory Analyses

We employed classical laboratory techniques for RNA extraction, cell culture, embryonated egg culture, microneutralization (MN). Detailed descriptions of these methods can be found in the Appendix.

#### Molecular Analysis

Extracted RNA specimens were screened with RT-qPCR for the influenza A virus (matrix gene) and for H5 influenza virus using WHO-recommended assays^13^. We also studied specimens by a conventional RT-PCR for coronaviruses^10,14^. Specimens with evidence of influenza A virus RNA were further characterized using conventional RT-PCR for the HA cleavage site according to Slomka *et. al*, 2007^15^. The 300-bp amplicons were visualized on a 1% agarose gel by electrophoresis. Amplicons of the size expected for the HA cleavage site protocol were sent for Sanger sequencing.

#### Next Generation Sequencing and Phylogenetic Analysis

RNA-seq library for samples selected for NGS was prepared using NEBNext Ultra II RNA Library Prep kit for Illumina (New England Biolabs, Ipswich, MA) following the manufacturer’s recommended procedure. The libraries were sequenced on the Illumina NextSeq 550 platform (Illumina Inc., San Diego, CA) for paired end 75 bp sequencing. Raw sequence reads were analyzed on the openly available CZ ID platform at (https://czid.org/)^16^, where host backgrounds were depleted, and taxon classification identified hits for influenza A viruses and reference-based consensus genomes were generated. The reads were also de novo assembled using SPADEs^17^ and abyss v2.3.7^18^ and contigs were blasted using NCBI blast against a custom made virus protein database and using NCBI blastn against NCBI nt database. Furthermore, reference-based reads assembly was carried out via Bowtie v1.1.2^19^. Multiple sequencing alignment of nucleotides was done using MAFFT^20^. IQ-tree v1.6.12^20^ was used to construct a maximum likelihood tree.

#### Microneutralization Assays (MN)

To determine if the farm workers had been exposed to the influenza A (H5N1) virus, we measured the neutralizing antibody to a recombinant H5N1 (rg-A/bald eagle/Florida/W22-134-OP/2022 of clade 2.3.4.4b) kindly provided by Dr Richard Webby of St. Jude Children Hospital, Memphis, TN using MN.

## Results

### Farm Information

We visited Farm A on April 3^rd^ and Farm B on April 4^th^, 2024. While we previously studied specimens from Farm A^21^, this was our first visit to the farm site. Farm A had 7,200 dairy cattle, was located on 4,900 acres of land, and employed 180 cattle workers. Farm B had 8,200 dairy cattle, was located on 98 acres of land, and employed 45 cattle workers. Farm A was solely engaged in dairy farming. Farm B raised both dairy and beef cattle, but dairy and beef cattle were kept in separate areas. No other livestock were raised on these farms. Additional information can be found in the appendix.

During the 30 days prior to our visit, both farms reported that their livestock had shown or were currently showing signs of respiratory disease including coughing, nasal discharges, difficulty breathing and fever. In addition, farm B reported receiving new stocks of animals each week but was unaware of any cattle illnesses reported from the sending farm. As recorded in our earlier report^21^ Farm A first noticed illnesses among cattle on March 6^th^. An estimate 4.75% of the herd was affected with illnesses largely waned by April 1st. Farm B first noted dairy cattle illnesses on March 20^th^ with the illnesses increasing over the next 13 days, eventually affecting an estimated 14% of the milking herd. On March 22, illnesses were first noted in the Farm B’s feral cats with cats showing lethargy, paralysis, and increased respiratory rate. Farm B estimated that 15-20 of their ∼40 feral cats died during the next 14 days (**Video**). Farm B thought they might have observed signs of illness in a beef cow which had recently calved during their dairy cattle epizootic but discounted same when a molecular test of the cow’s sera was negative for influenza A RNA. As measured by milk production the epizootic quickly abated and Farm B stopped segregating sick cows on April 6^th^.

### Farm Worker Demographics

By our previously IRB-approved protocol we were permitted to enroll up to 10 animal workers on each farm. In total, we enrolled via informed consent 17 farm workers (10 on Farm A and 7 on Farm B). Twelve farm workers were male (70.6%, **Table 1**). In Farm A, a 100% permitted both nasopharyngeal (NP) swab and serum collection. On farm B, all permitted NP swab collections but only 4 agreed to serum collection. A majority of the farm workers (88.2%) were of Latino ethnicity (**Table 1**). Five (29.4%) of the 17 farm workers reported having experienced recent respiratory illnesses and using different medications including antibiotics, ibuprofen, multivitamin, and cough syrup in the last 30 days (**Table 1**).

**Table 1:**
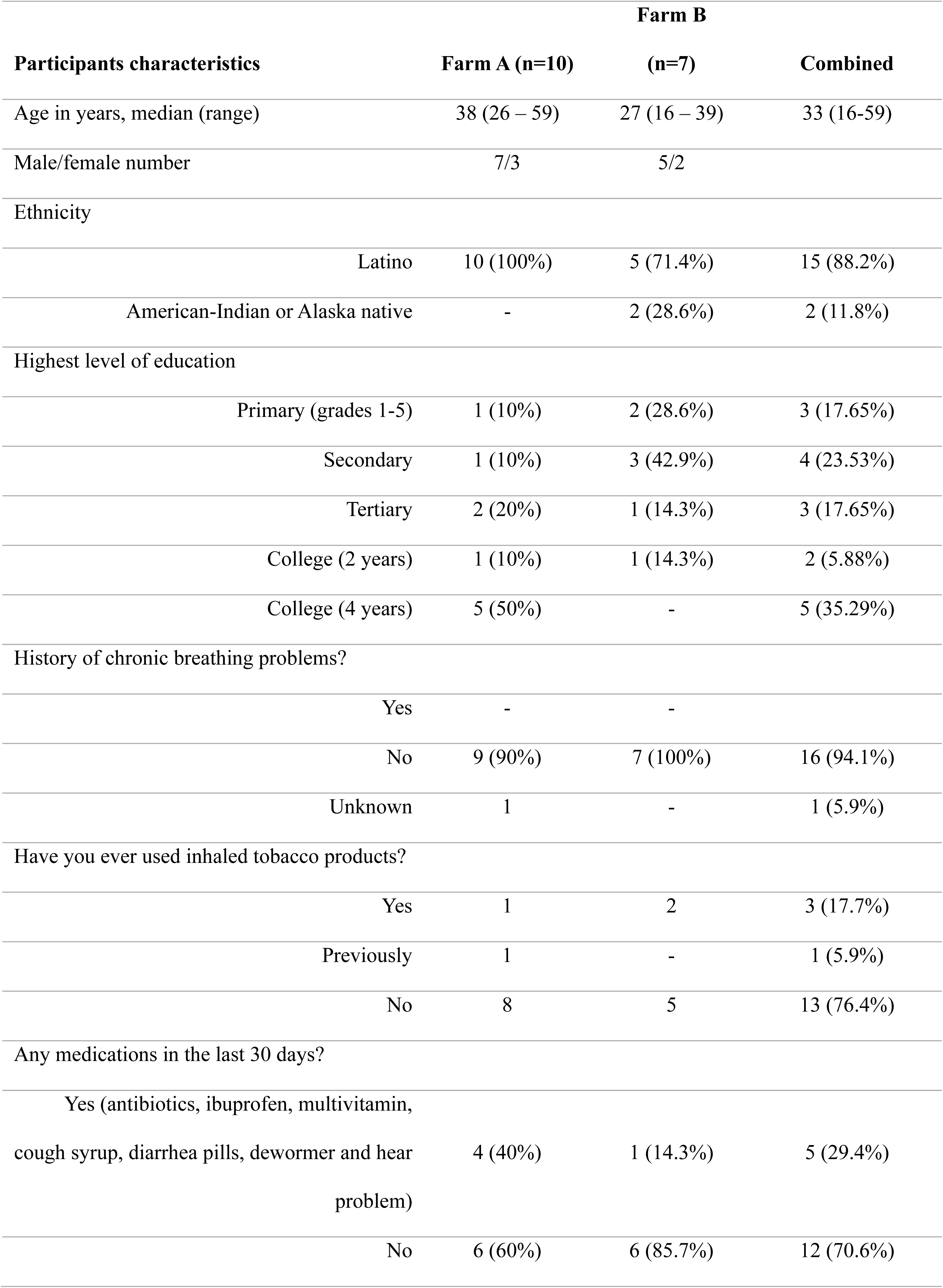
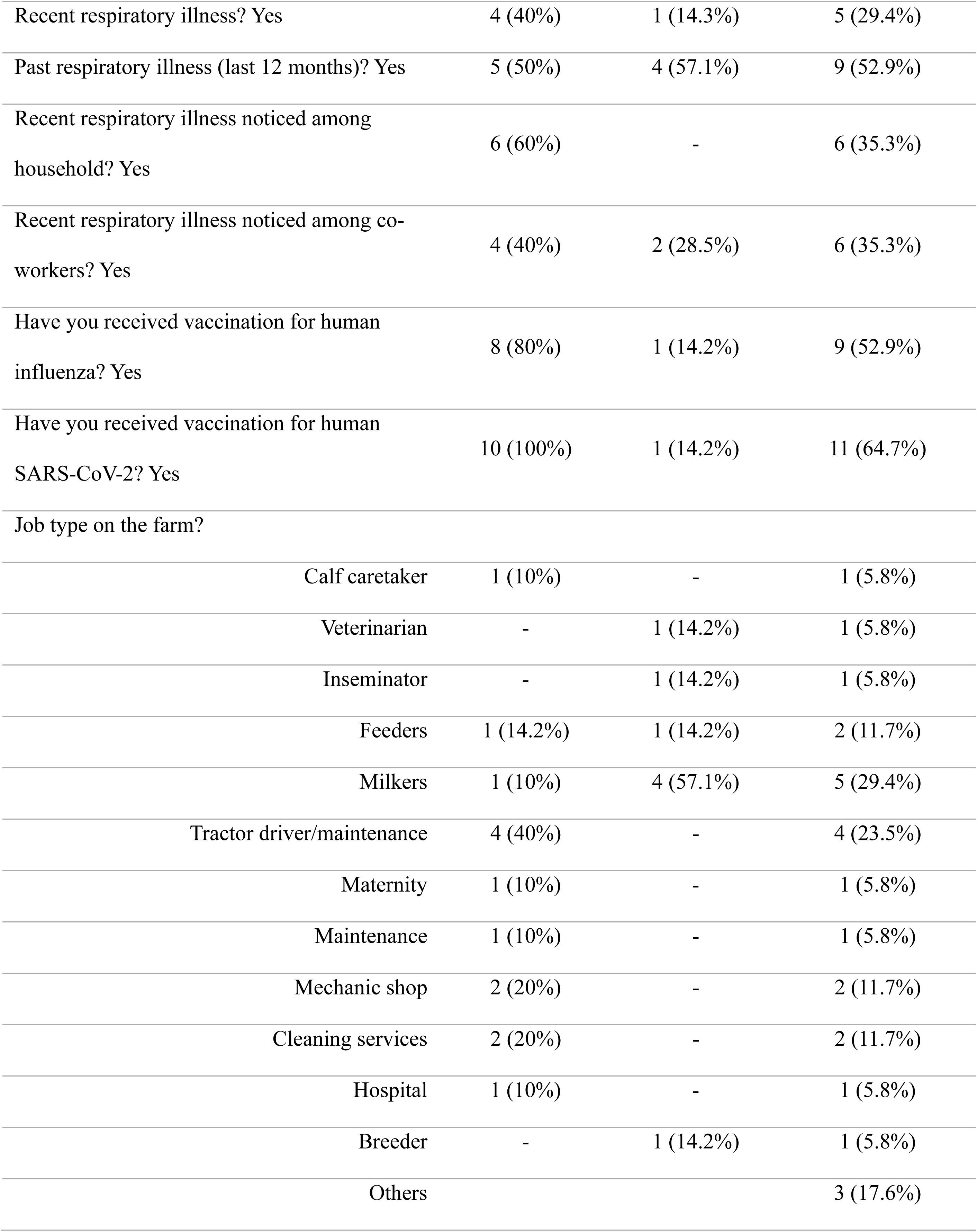
Demographic characteristics of participants from the two dairy farms.

### Laboratory Studies

#### Human Samples

All 17 NP swabs collected from farm workers were negative by molecular assays for influenza A viruses and coronaviruses. Microneutralization assays (MN) conducted on the fourteen farm workers’ sera samples indicated a prevalence of 14.3% (2/14) of neutralizing antibodies to a recombinant influenza A H5N1 virus. All the MN positive samples (20%, 2/10) were from Farm A (**Table 2**).

**Table 2.** Farm workers clinical history and laboratory assay results.

| Sample ID |  | Respiratory symptoms | Nasopharyngeal swab |  | MN* |
| --- | --- | --- | --- | --- | --- |
| number | Farm | during the last 12 months? | influenza A qRT-PCR (Ct) | MN* titer | interpretation |
| USH_01 | Farm A | No | Negative | 1:20 | Negative |
| USH_02 | Farm A | Yes | Negative | 1:10 | Negative |
| USH_03 | Farm A | Yes | Negative | 1:10 | Negative |
| USH_04 | Farm A | Yes | Negative | <1:10 | Negative |
| USH_05 | Farm A | No | Negative | **1:40** | **Positive** |
| USH_06 | Farm A | Yes | Negative | 1:20 | Negative |
| USH_07 | Farm A | No | Negative | 1:20 | Negative |
| USH_08 | Farm A | Yes | Negative | 1:20 | Negative |
| USH_09 | Farm A | Yes | Negative | 1:20 | Negative |
| USH_10 | Farm A | Yes | Negative | **1:80** | **Positive** |
| USH_11 | Farm B | No | Negative | 1:20 | Negative |
| USH_12 | Farm B | No | Negative | 1:20 | Negative |
| USH_13 | Farm B | Yes | Negative | NA | NA |
| USH_14 | Farm B | Yes | Negative | NA | NA |
| USH_15 | Farm B | No | Negative | 1:20 | Negative |
| USH_16 | Farm B | Yes | Negative | 1:20 | Negative |
| USH_17 | Farm B | Yes | Negative | NA | NA |

\*Serologic microneutralization (MN) assays were run against recombinant H5N1 (rg-A/bald eagle/Florida/W22-134-
OP/2022 of clade 2.3.4.4b), NA = Not applicable

#### Animal Samples

In all, 39 deep nasal swabs were collected from cattle on the farms. Among these only nasal swab specimen (USL_042 collected from Farm B) had molecular evidence of influenza A virus by the RT-qPCR assay (**Table 3**). This specimen was obtained from a recuperating cow and had a relatively high Ct value of 38.76. The virus was successfully isolated and propagated in embryonated eggs (but not in MDCK or MDBK cells). It was characterized as HPAIV H5N1. In addition, multiple other specimens collected at Farm B were positive by RT-qPCR for influenza A virus and successfully grown in embryonated eggs or two different cell lines (**Table 3**). These included 9 (64%) of 14 milk samples (Ct value from 22.26 to 29.88). Most had molecular evidence of HPAIV H5N1 virus. In addition, one cattle swab specimen (USL_022) had evidence of a coronavirus by conventional RT-PCR having bands of correct molecular weight. Sanger sequencing revealed the cow had evidence of SAR-CoV-2 infection.

**Table 3.**
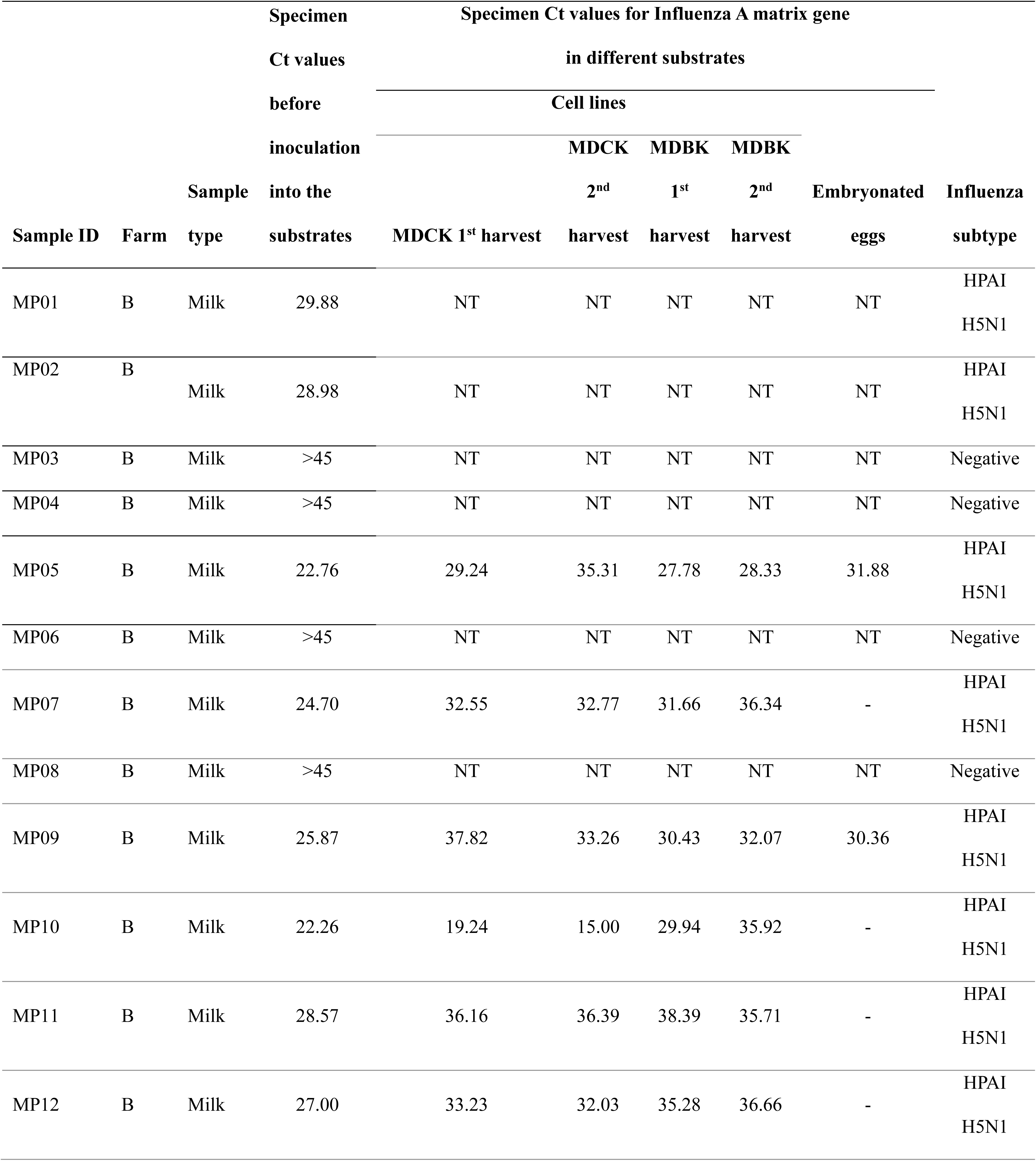
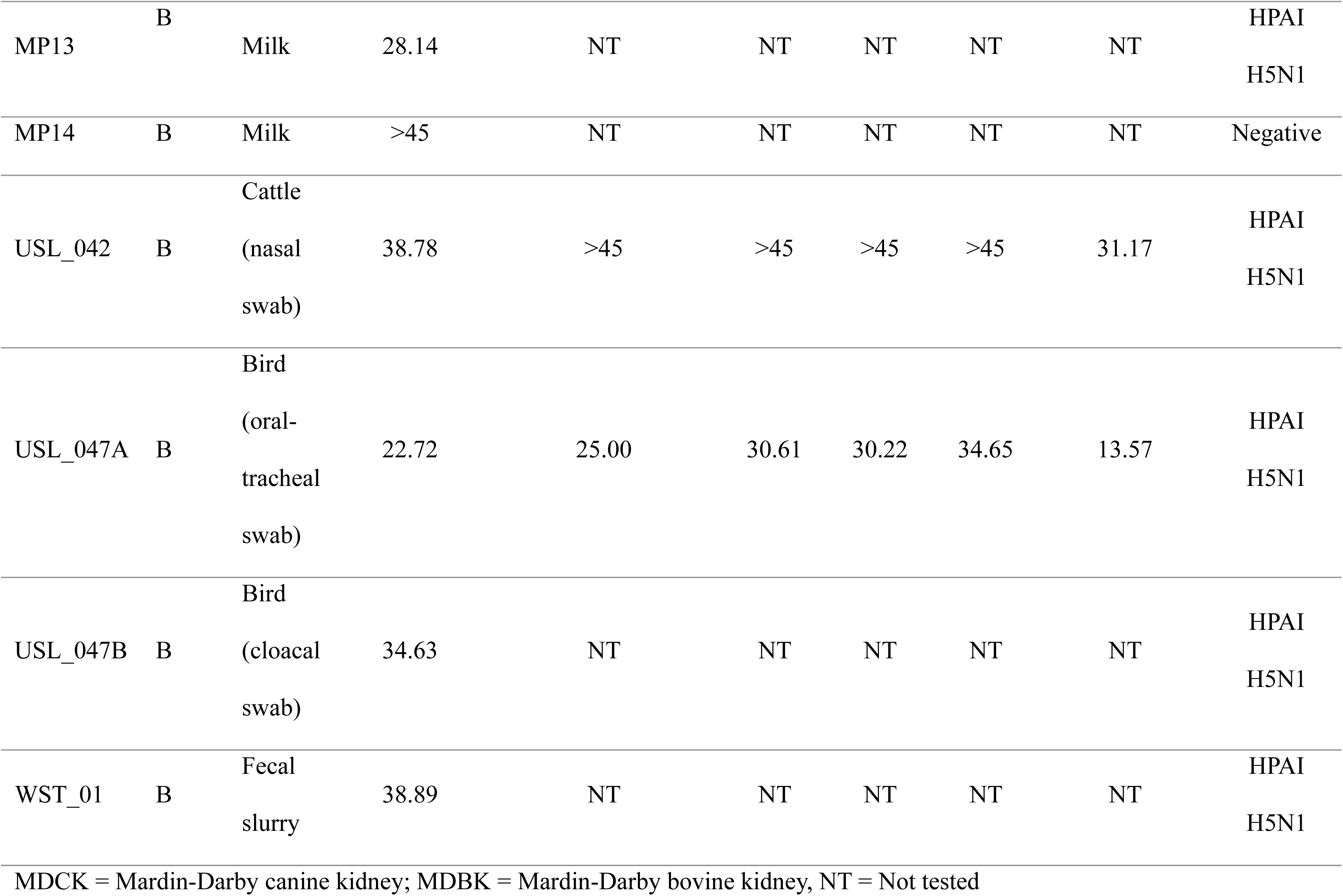
Comparison of the threshold cycle (Ct) of the different sample types before and after inoculation of cell lines and embryonated eggs.

Oropharyngeal and cloacal swab specimens were obtained from a dead female great-tailed grackle (*Quiscalus mexicanus*) (**Fig. 1**) found in an open-air dairy cattle barn. Both swabs had molecular evidence of influenza A virus (matrix gene) and that were characterized as HPAIV H5N1 (**Table 3**).

**Fig. 1.**
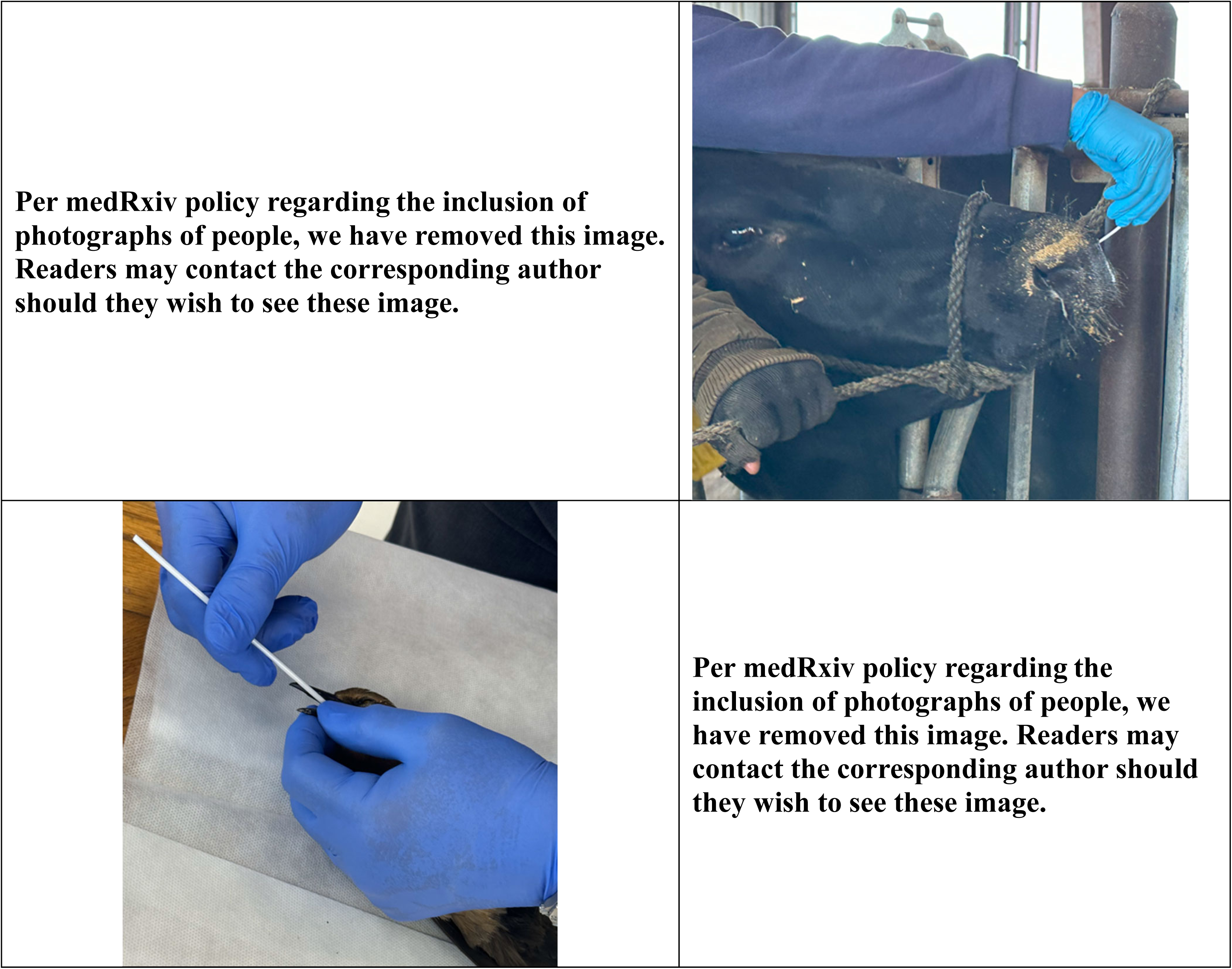
Images from various farm visits demonstrating sample collections. Nasal swabs were taken from sick and healthy cattle, oral-tracheal and cloacal swabs were taken from a dead bird, and aerosol samples were taken from cattle environments.

#### Environmental Samples

Due to the TE-BC251 NIOSH bioaerosol cyclone samplers (Tisch Environmental Inc., Cleves, OH) having three collection chambers, the four bioaerosol samplers we employed on each farm yielded 12 samples each. None of these 24 samples had molecular evidence of influenza A virus or coronaviruses. A single cattle fecal slurry sample was collected on Farm B and it had molecular evidence of influenza A virus by qRT-PCR with a high Ct value (38.89) (**Table 3**).

### Sanger Sequencing of Influenza A HA cleavage site

In studying the specimens with evidence of influenza A virus, conventional RT-PCR studies of HA and NA genes yielded good quality Sanger sequencing results from the dead bird’s oral-tracheal swab sample and from six milk samples (all from Farm B). For phylogenetic analysis, we used sequences from these seven samples. The phylogenetic analyses showed all seven sequences were within the same clusters implying they were closely related and from the same location. Furthermore, our sequences were in the same HA clade as those of other Texas sequences deposited in GenBank that were obtained during the current epizootic. Sanger sequencing results for the six milk (and the dead bird) samples demonstrated the presence of multiple basic amino acids (PLREKRRKRGLF) at the HA cleavage site indicating that bovine strains were highly pathogenic avian influenza (HPAI) H5N1 viruses belonging to clade 2.3.4.4b. (**Fig. S1 in Appendix**)

### Next Generation Sequencing Results

We submitted five samples for NGS: the cattle nasal swab with molecular evidence of SARS-CoV-2, one cow nasal swab specimen, two milk specimens, and the oral-tracheal swab specimen from the dead grackle. The cattle nasal swab specimens with SARS-Cov-2 did not pass library prep quality control. Maximum likelihood phylogenetic analysis of HA (**Fig. 2**) and NA genes (**Fig. 3**) from the other four specimens revealed that their genomes clustered with H5N1 influenza A virus HA clade 2.3.4.4b in GenBank. The nucleotide sequences from the dairy cow and milk samples shared 100% identity scores with the genome from the dead grackle. The milk and swab samples shared 99.94% identity, while the dead bird and milk samples shared pairwise identity ranging 99.94 to 100%.

**Fig. 2.**
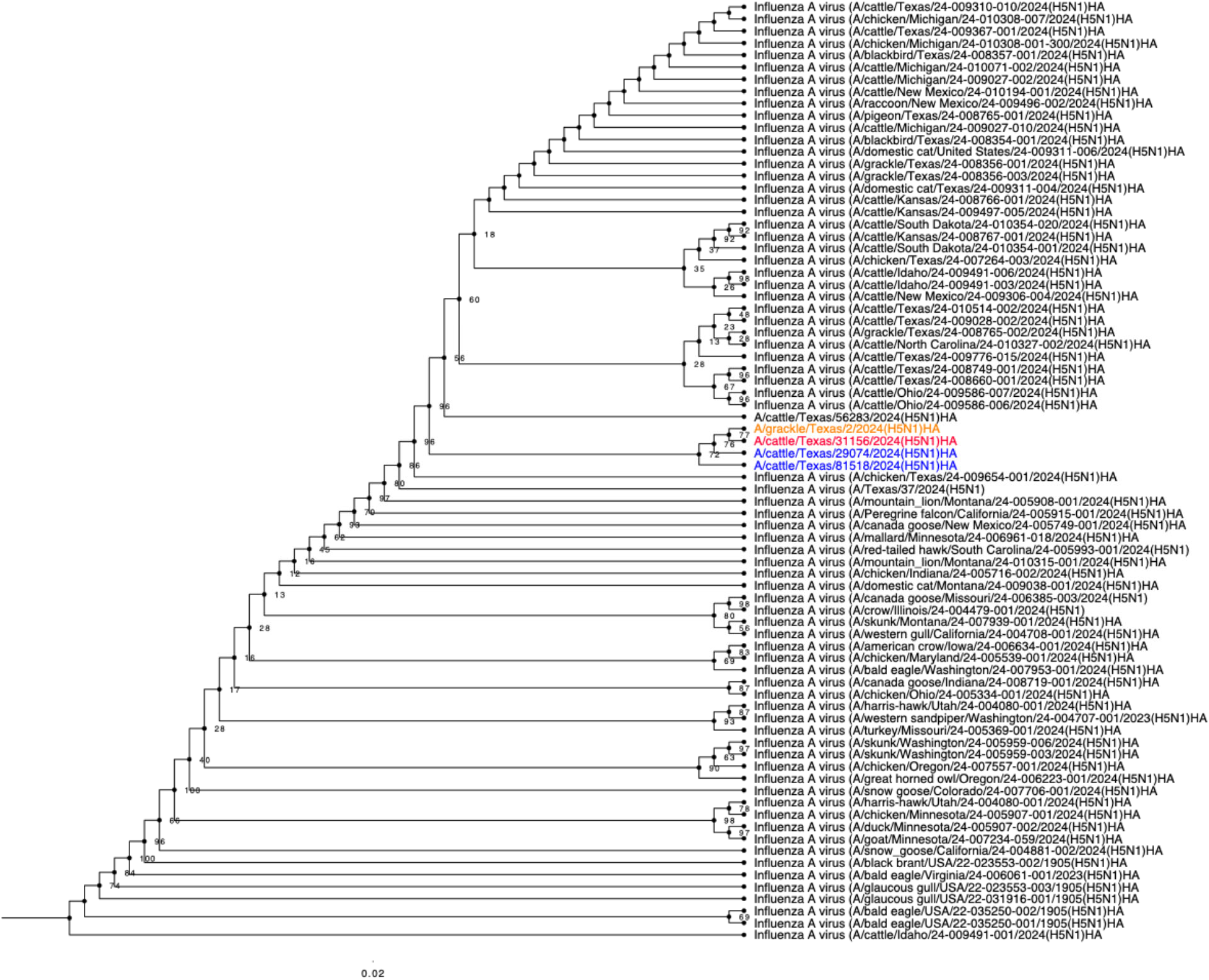
Phylogenetic Tree of the HA gene. Maximum likelihood phylogenetic tree inferred for four viruses isolated in this study (two from milk-colored blue, one from a nasal swab of cow-colored red, and one from a dead grackle colored orange), to other related HPAI H5N1 viruses submitted to GenBank during the current epizootic, downloaded from NCBI. Bootstrap values are displayed at key nodes.

**Fig. 3.**
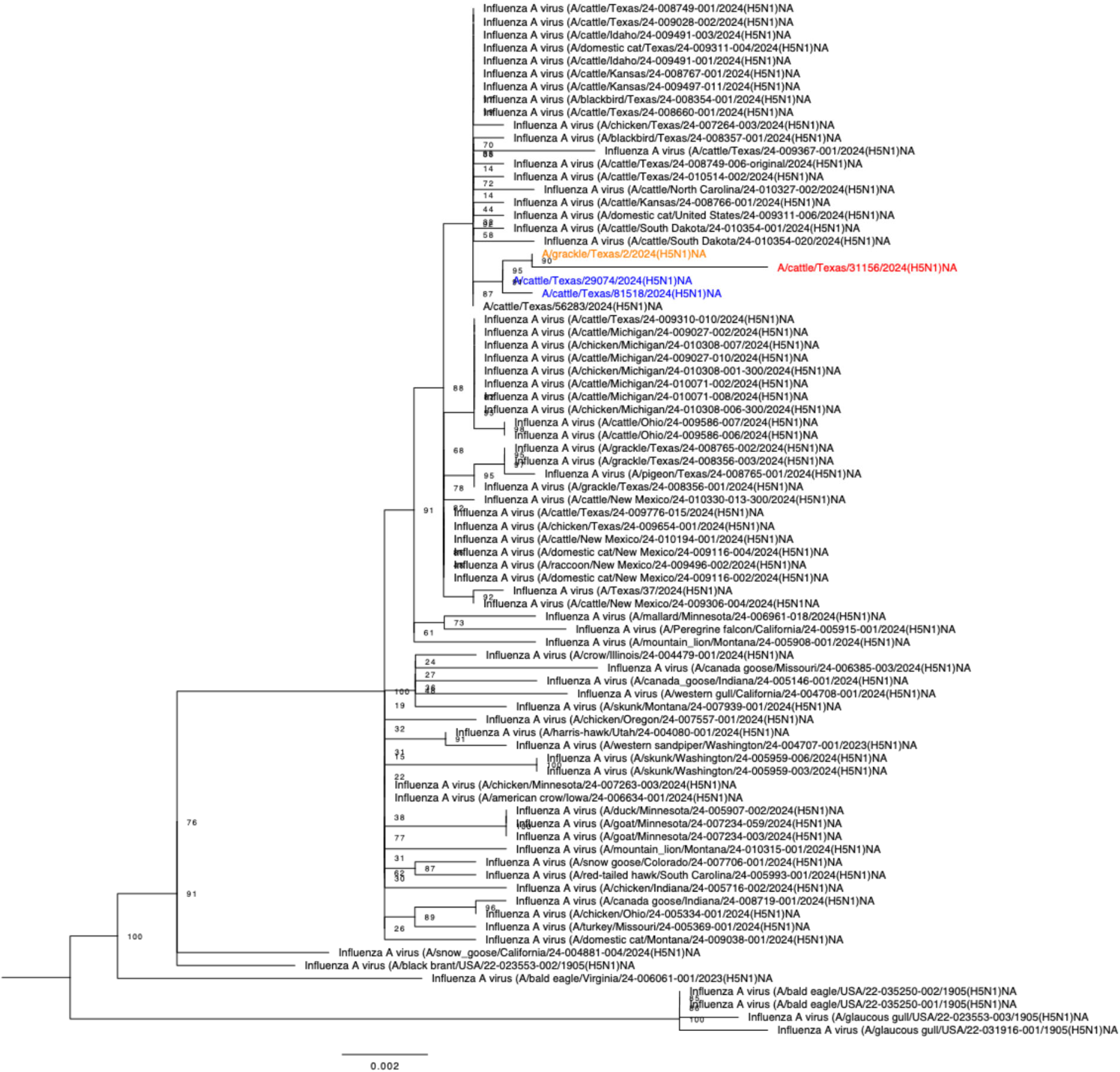
Phylogenetic Tree of the NA gene. Maximum likelihood phylogenetic tree inferred for four viruses isolated in this study (two from milk-colored blue, one from a nasal swab of cow-colored red, and one from a dead grackle colored orange), to other related HPAI H5N1 viruses submitted to GenBank during the current epizootic, downloaded from NCBI. Bootstrap values are displayed at key nodes.

### Mutation Analysis

Several common mutations were identified across the four viral genomes reported in this study (**Table S1 in Appendix**). We identified several mutations that alter host cell specificity, target drug binding sites and known to cause antigenic shifts or cause mild drug resistance. These types of mutation are assigned with a level 2 warning/significance or orange mutations according to FluSurver (http://flusurver.bii.a-star.edu.sg/). Across the various segments of the four genomes, we found several mutations associated with viral virulence and host specificity shifts. Virulence-based mutations were detected in *PB2* (V495I and M676A) and *PB1* (N375S) gene segments. Mutations associated with host specificity shift included two in the *HA* gene (N110S and V226A). A rather infrequent mutation was found in the PB2 gene (M631L) of all four viruses. This mutation increases H5N1 HPAIV propagation in human cells by enhancing polymerase activity and virus replication.

## Discussion

The US H5N1 epizootic is unprecedented for its rapid spread in the United States and impact upon numerous wild and livestock animal species. While our field sampling was limited, this report is important in adding new observations. It seems likely that H5N1 HPAIV detections in the nasal passage of cattle occur early in infection and are brief in duration. Workers in Farm A first noticed cattle illness on March 6^th^ and by April 1^st^ cattle illnesses had largely waned. We found H5N1 HPAIV virus in 6 (42%) of 14 sick cattle nasal swabs on March 21^st^, 1 (10%) of 10 sick cattle nasal swabs on April 1^st^, and none of 14 sick cattle nasal swabs on April 3^rd^. On Farm B, cattle illnesses were first observed on March 20th. When we visited the farm on April 4^th^, we found only 1 (7%) of 14 ill cows to have evidence of H5N1 HPAIV in their nasal swabs. While the virus seems to rapidly clear from cattle nasal tissues, infected cows may shed H5N1 for longer periods in their milk. Further research will be required to establish the duration of H5N1 shedding in infected cow’s milk. On Farm B, while nasal swabs had little evidence of H5N1 HPAIV when we visited on April 4^th^, 9 (64%) of 14 cattle milk samples from convalescing cows had evidence of virus with relatively low Ct values (high titers of virus). While our sampling were not likely representative of the many cows on the farms, the available data seems consistent with reports from US Department of Agriculture^22,23^ and cattle associations^24^ that HPAIV H5N1 affects only a subset of cattle and the generally moderate illnesses quickly resolve.

Additionally, in one of the only serological studies of dairy workers during this epizootic, two (20%) of 10 dairy workers from Farm A who donated sera had evidence of elevated titers against a recombinant H5N1 virus of clade 2.3.4.4b virus by a MN assay. The first of these dairy workers had a moderately elevated MN titer of 1:40. He often worked inside cattle corrals close to dairy cattle. He reported no respiratory illnesses during the last 12 months but reported having a cough and taking cough medication at the time we enrolled him. The second worker had a MN of 1:80. She worked in the Farm A’s cafeteria. She reported experiencing fever, cough or sore throat during that last 12 months as well as being around others at work with similar respiratory signs and symptoms. She had just recovered from a respiratory illness when we enrolled her.

While we cannot rule out cross-reacting antibodies from previous influenza A virus infections or vaccines as a cause for the MN titer elevations, neutralizing assays are often considered the best assay for the virus-specific serological assessments. We observe that workers in Farm A had more time (∼4 weeks) to develop antibodies to the H5N1 virus as compared to workers on Farm B (∼2 weeks) as Farm A experienced the H5N1 epizootic 14 days earlier than Farm B.

The study is also unique in documenting a likely SARS-CoV-2 infection in the nasal secretions of a sick cow. SARS-CoV-2 infections in cattle is known but thought to be rare^25,26^. A cross-sectional study of plasma from 1000 cattle from 83 farms in Germany^27^ during 2021-22 suggested that 11 cattle from 9 farms or ∼1% of cows tested had evidence of previous infection. Experimental data also suggest that the risk of SARS-CoV-2 infection in cattle is low^28^. Our report is important in that the observation by Farm B’s manager that they thought they had detected H5N1 HPAI in a beef cow that had just delivered is concerning. Farm B discounted their hunch when a molecular assay from the cow’s blood was negative for influenza A. This hunch may have been correct in that the time period of cattle viremia may be relatively short. We now know that a better choice would have been a molecular assay of the cow’s milk.

In addition, this report is valuable in corroborating reports of both dead cats and birds that were observed associated with disease cattle in Farm B. Observing feral cat and bird die-offs is likely a tell-tale sign of HPAI incursions on farms. Our study is further valuable for the identification of numerous mutations associated with viral spillover potential. Finally, it seems important to note the H5N1 HPAIV isolates from cattle, cattle milk, and the dead bird in this study cluster closely with other H5N1 HPAIV associated with this now national epizootic.

Continued surveillance and reporting of results from deidentified farms/workers is important in understanding outbreak trends. Genome analyses of future H5N1 strains is also extremely important, not only for determining which viral strains are circulating, but also in assessing genetic markers associated increased virulence and resistance to antivirals. Finally, information from this type of surveillance work is helpful in directing vaccine production and in considering employment of vaccines to prevent human disease, livestock disease or both.

It now seems especially prudent that we find ways to prospectively and more intensely study dairy farms to better quantify serological evidence of infections in both livestock and dairy workers. Before we can perform such important research, we need to find ways to fully protect the dairy businesses from any economic harm that might arise through such intensive study.

## Acknowledgments

This project was supported in part by the Agriculture and Food Research Initiative Competitive Grant from the American Rescue Plan Act (award number 2023-70432-39558) through USDA APHIS and Professor Gregory C. Gray’s startup funding from the University of Texas Medical Branch. The findings and conclusions in this presentation are those of the authors and should not be construed to represent any official USDA or US Government determination or policy. We thank Dr. Richard Webby of St. Jude Children Hospital, Memphis, TN for sharing the recombinant H5N1 (rg-A/bald eagle/Florida/W22-134-OP/2022) virus used in the MN assays.

We thank Barbara Petersen, DVM of Sunrise Veterinary Service for her education regarding livestock farming. We thank the dairy farm owners and managers for engaging us in research collaboration. We gratefully acknowledge all data contributors, i.e., the authors and their originating laboratories responsible for obtaining the specimens, and their submitting laboratories for generating the genetic sequence and metadata and sharing via NCBI the, on which this research is based.

## Author Contributions

Conceptualization: GCG

Methodology: IS, LVM, JUO, LVM, JAL, CMTV, ALM, HH, GCG

Investigation: IS, DS, JUO, LVM, GGO, JAL, NES, HH, GCG

Data analysis: IS, JUO, LVM, HH

Visualization: JUO, LVM

Funding acquisition: GCG

Project administration: GCG, DS, LVM

Supervision: GCG, LVM

Writing – original draft: IS, GCG

Writing – review & editing: IS, DS, JUO, LVM, JAL, CMJV, GCG

## Data Availability

Data needed to evaluate the conclusions in the paper are present in the paper and/or the supplementary materials. All viral sequences have been deposited on NCBI with accession numbers: PP914075-PP914106. Researchers with BSL3Ag-approved laboratories may request the live viruses A/cattle/Texas/81518/2024(H5N1) and A/cattle/Texas/29074/2024(H5N1) isolated from milk and A/grackle/Texas/2/2024(H5N1) isolated from dead female great-tailed grackle (*Quiscalus mexicanus*) described in this paper by contacting Dr. Kenneth Plante, PhD of UTMB’s World Reference Center for Emerging Viruses and Arboviruses (https://www.utmb.edu/wrceva/). Additional data, study instruments, or specimens may be requested from the corresponding author. Farm location, farm identification and personal identifying data for human are protected by nondisclosure agreement and will not be shared. The sharing of specimens or data will require the signing of material transfer agreement (MTA).

## Video

Video of sick feral cat taken on Farm B during the HPAI H5N1 incursion (download from link). https://liveutmb-my.sharepoint.com/:v:/g/personal/gcgray_utmb_edu/EXe2BTggZo9HhE84hi-G0sUBdpRGQDqFVim-pWHSAbSHsA?nav=eyJyZWZlcnJhbEluZm8iOnsicmVmZXJyYWxBcHAiOiJPbmVEcml2ZUZvckJ1c2luZXNzIiwicmVmZXJyYWxBcHBQbGF0Zm9ybSI6IldlYiIsInJlZmVycmFsTW9kZSI6InZpZXciLCJyZWZlcnJhbFZpZXciOiJNeUZpbGVzTGlua0NvcHkifX0&e=fJxEdR

## Supplemental Appendix

### Additional Text

#### Methods

##### Sample Preparation

*N*asopharyngeal (NP) swabs were collected using human volunteers a 6-inch polyester-tipped applicator swab (Thermo Fisher Scientific, Pittsburgh, PA). After collection, swabs were broken off into a 15mL tube containing 2mL of viral transport medium (VTM; rmbio, Missoula, MT) containing Hank’s basal salt solution, fetal bovine serum, gentamicin and amphotericin B and kept in an insulated portable cooler before being transported to the One Health Laboratory at UTMB, Galveston. At the laboratory, the VTM samples were vortexed and aliquoted before being stored at –80°C. In addition, approximately 10mL of whole blood was collected from consenting farm workers. The blood was allowed to clot and kept in an insulated portable cooler. Thereafter, the blood was centrifuged at 1300 rpm for 15 minutes. Aliquots of the serum specimens were made and stored at –80°C until studied with the microneutralization assay.

Animal samples were collected by farm staff. They selected 14 sick animals and six healthy animals per farm. Samples were placed in 2ml of VTM and kept cool before being transported to UTMB and preserved at –80°C until tested.

To examine the hypothesis that novel viruses might be detected in aerosol on the farms, we used National Institute for Occupational Safety and Health (NIOSH) bioaerosol cyclone samplers (Tisch Environmental Inc., Cleves, OH) as previously reported^1^. Prior to set up, each NIOSH sampler was calibrated to a flow rate of 3.5 L/min^1^, this flow rate was obtained using an Air Check Touch (part number: 220-5000TC) pump from SKC INC. In each farm, research staff placed four bioaerosol samplers were set up in different farm locations where cattle and farm staff mixed. The bioaerosol samplers were run for ∼3 hours, then removed and kept in a refrigerated cooler. The locations and times the samplers were started and stopped were documented. Each tube and the filter attached to the samplers were hydrated with 1 mL of 0.5% protease-free bovine serum albumin (w/v) (ThermoFisher Scientific, Waltham, MA cat no. BP9703100) in phosphate buffered saline, vortexed and aliquoted. The aliquots were stored at – 80°C until analyzed.

##### RNA Extraction

On the QIAcube Connect automated extraction system (Qiagen) using the QIAamp Viral RNA Mini Kit (Qiagen, Valencia, CA), RNA was extracted from 140µl aliquots of specimens (nasal swabs, milk and bioaerosol including waste from the tank) collected from humans, animals and the environment following the manufacturer’s instructions. RNA was eluted with 60µl of buffer AVE and stored at –80°C until tested.

##### Cell Culture

To isolate virus, Madin-Darby canine kidney (MDCK) (ATCC, cat no. CRL-CCL34) and Madin–Darby bovine kidney (MDBK) (ATCC, cat no. CCL-22) cell lines were grown in Minimal Essential Medium (MEM) containing Earles salt and L-glutamine (ThermoFisher Scientific, Waltham, MA cat no. 11095080) supplemented with 10% fetal bovine serum (ThermoFisher Scientific, Waltham, MA cat no. 26140-079) and 1X penicillin-streptomycin. The cells were cultured as monolayers in 6-well plates at 37°C in 5% CO_2_ environment.

Prior to infecting the cells, all specimens were filtered using sterile 0.45uM pore-size filters (Millipore Sigma™, Ireland cat no. SLGVR33RS). At confluency, the cells were washed with PBS (Corning, Manassas, VA) and infected with 0.2ml of the filtered specimens with an addition of 0.8 ml of serum-free maintenance medium (MEM, 1X penicillin-streptomycin and 2 µg/mL TPCK-Trypsin).

##### Embryonated Egg *Culture*

Ten-day-old specific-pathogen-free (SPF) chicken eggs (AVS Bio, Norwich, CT) were used for growing AIV isolates from RT-qPCR influenza virus RNA-positive specimens (1 nasal swab from cattle, 6 samples of milk and fecal and oral swab from dead bird) according to a standard protocol^2^. Briefly, SPF eggs were incubated for 10 days at 37°C and 45% humidity. The eggs were monitored daily using an egg candling lamp. All specimens were filtered using 0.45uM pore-size filters (Millipore Sigma) before inoculation with 0.2ml of the filtrate into the allantoic cavity of the eggs.

The inoculated eggs were candled daily to check for embryo death. After the incubation period, eggs were chilled at 4°C overnight. Allantoic fluid from the dead eggs were then harvested and frozen at –80°C until tested. The virus work was conducted in our BSL3Ag laboratory. For molecular analyses the harvested allantoic fluid was treated with TRizol LS Reagent (Invitrogen, Waltham, MA) under BSL3Ag conditions before being moved into BSL2 where they underwent RNA extraction following the manufacturer’s recommendations. RNA was then stored at –80°C for further molecular testing.

##### Microneutralization Assays (MN)

We measured the neutralizing antibody to a recombinant H5N1 (rg-A/bald eagle/Florida/W22-134-OP/2022 of clade 2.3.4.4b) kindly provided by Dr Richard Webby of St. Jude Children Hospital, Memphis, TN using previously described MN procedures^3^. Prior to testing, sera were treated overnight (18-24 hours) with receptor destroying enzyme (RDE, Denka Seiken, Japan) according to the manufacturer’s instructions to cleave sialic acid from glycoproteins and glycolipids in the serum, destroying these potential inhibitors from interfering with the HAI assay, and subsequently heat inactivated at 56°C for 30 min. The RDE-treated sera were then diluted with PBS to a final dilution of 1:10 as described by Cuevas et al^3^. Two-fold dilutions of the serum starting with the 1:10 dilution were performed. We considered a titer ≥ 1:40 as positive.

The recombinant H5N1 virus was propagated and titrated to determine the 50% tissue culture infectious dose (TCID50) in MDCK cells^4^. The end point titer of the recombinant H5N1 was calculated according to the Spearman-Karber formula^5^. The MN was performed in 96-well microtiter plates with the inactivated sera and the recombinant H5N1 according to a published protocol^3^.Each of the samples were tested in duplicate and the lowest serum dilution without CPE was recorded as the neutralizing titer.

### Results

#### Farm Information

Neither farm had provisions for preventing wild birds from accessing cattle areas. The cattle feed or water troughs were open to birds. Both farms reported periodically vaccinating dairy cattle with vaccines against common cattle respiratory pathogens. Each farm reported using numerous biosecurity measures including cleaning and disinfection of clothing and equipment as well as segregation of sick animals. The workers in Farm A reported always being provided with personal protective equipment (PPE) including aprons, face shield, disposable gloves, washable boots or disposable booties and sometimes mask and frequent handwashing. Workers on Farm B reported always being provided with coveralls, aprons, disposable latex gloves, washable boots and frequently washing their hands. They reported only occasionally using eye protection glasses and rarely using masks.

**Fig. S1.**
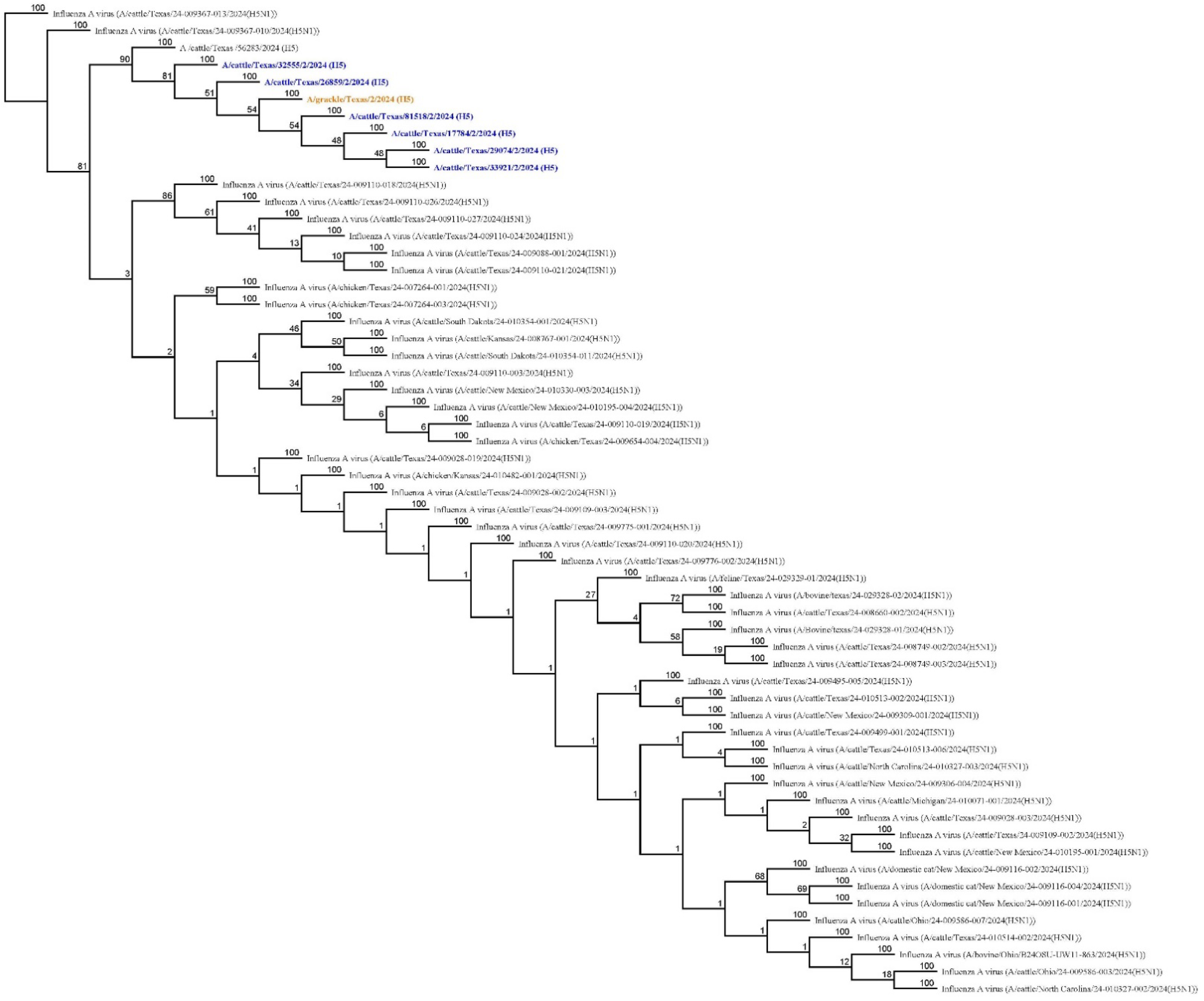
Phylogenetic Tree of the HA cleavage site. Neighbor-joining phylogenetic tree of the HA cleavage sites from six cattle milk sample (colored blue) sequences and one dead grackle (colored orange) amplified from this study compared to other related viruses in GenBank from NCBI. Phylogenetic analyses were performed using the Geneious Prime software v2024.0.5.

**Table S1.** Summary of identified mutations. As shown in FluSurver (http://flusurver.bii.a-star.edu.sg). Mutations that alter host-cell specificity or occur at drug-binding sites are labelled orange mutations and assigned a level 2 significance warning. This category also includes mutations that cause antigenic shifts or moderate drug resistance.

| Sample ID | Mutation Region | All mutations | Orange Mutations | Reference |
| --- | --- | --- | --- | --- |
| A/CATTLE/TEXAS /29074/2024(H5N1) HA | HA | K3N;G16S;N110S;L120M;L131Q;T139P;T156A;Q185R;V194I;A201E;T211I;V226A;N252D;E284G;M285V;I298V;K492E;I526V;V538A;I547M;V548M | N110S;L131Q;T139P;V226A | HA A/Sichuan/26221/2014(H5N6) |
| A/CATTLE/TEXAS /29074/2024(H5N1) MP | M1 | N82S;N85S;N87T;T140A;F144L;M165I;V166A;A200V;A227T;K230R;M248L |  | M1 A/Duck/Guangdong /E1/2012(H10N8) |
| A/CATTLE/TEXAS /29074/2024(H5N1) MP | M2 | R12K;K18N;I51V;R61G;D88N |  | M2 A/Mallard/Astrakhan/263/1982(H14N5) |
| A/CATTLE/TEXAS /29074/2024(H5N1) NA | NA | I8T;V17I;I20V;H44Y;A46P;V67I;N71S;T76A;K78Q;A81T;V99I;H100Y;H155Y;T188I;M258I;L269M;E287D;T289M;V321I;G336S;V338M;S339P;P340S;N366S;G382E;A395E;I418M;S434N;D460S |  | NA A/Goose/Guangdong/1/1996(H5N1) |
| A/CATTLE/TEXAS /29074/2024(H5N1) NS | NS1 | S7L;R21Q;S87P;C116S;D139N;A223E;V226I | A223E;V226I | NS1 A/Duck/Guangdong /E1/2012(H10N8) |
| A/CATTLE/TEXAS /29074/2024(H5N1) NS | NS2 | R61K;E67G | E67G | NS2 A/Duck/Guangdong /E1/2012(H10N8) |
| A/CATTLE/TEXAS /29074/2024(H5N1) NP | NP | Y52H;S482N |  | NP A/Chicken/BCFAV8 //2014(H5N2) |
| A/CATTLE/TEXAS /29074/2024(H5N1) PA | PA | I61M;T85A;K113R;L219I;S277P;M441V;K497R;Y535H;I543L;S558L;T608S |  | PA A/Netherlands/219/2003(H7N7) |
| A/CATTLE/TEXAS /29074/2024(H5N1) PB1 | PB1 | T59S;E75D;M171V;V179I;N375S;R430E;A587P | N375S | PB1 A/Mallard/Astrakhan/263/1982(H14N5) |
| A/CATTLE/TEXAS /29074/2024(H5N1) PB2 | PB2 | T58A;V109I;V139I;E362G;K389R;D441N;V478I;V495I;M631L;V649I;M676A | V495I;M676A |  |
| A/CATTLE/TEXAS /81518/2024(H5N1) HA | HA | K3N;G16S;N110S;L120M;L131Q;T139P;T156A;Q185R;V194I;A201E;T211I;V226A;N252D;E284G;M285V;I298V;K492E;I526V;V538A;I547M;V548M | N110S;L131Q;T139P;V226A | HA A/Sichuan/26221/2014(H5N6) |
| A/CATTLE/TEXAS /81518/2024(H5N1) M2 | M1 | N82S;N85S;N87T;T140A;F144L;M165I;V166A;A200V;A227T;K230R;M248L |  | M1 A/Duck/Guangdong /E1/2012(H10N8) |
| A/CATTLE/TEXAS<br>/81518/2024(H5N1)<br>M2 | M2 | R12K;K18N;I51V;R61G;D88N |  | M2<br>A/Mallard/Astrakha<br>n/263/1982(H14N5) |
| A/CATTLE/TEXAS<br>/81518/2024(H5N1)<br>NA | NA | I8T;V17I;I20V;H44Y;A46P;V67I;N71S;T76A;K78Q;A81T;V99I;H100Y;H155Y;T188I;M258I;L269M;E287D;T289M;V321I;G336S;V338M;S339P;P340S;N366S;G382E;A395E;I418M;S434N;D460G |  | NA<br>A/Goose/Guangdon<br>g/1/1996(H5N1) |
| A/CATTLE/TEXAS<br>/81518/2024(H5N1)<br>NEP | NS1 | S7L;R21Q;S87P;C116S;D139N;A223E;V226I | A223E;V226I | NS1<br>A/Duck/Guangdong<br>/E1/2012(H10N8) |
| A/CATTLE/TEXAS<br>/81518/2024(H5N1)<br>NEP | NS2 | R61K;E67G | E67G | NS2<br>A/Duck/Guangdong<br>/E1/2012(H10N8) |
| A/CATTLE/TEXAS<br>/81518/2024(H5N1)<br>NP | NP | Y52H;S482N |  | NP<br>A/Chicken/BCFAV8<br>//2014(H5N2) |
| A/CATTLE/TEXAS<br>/81518/2024(H5N1)<br>PA | PA | I61M;T85A;K113R;L219I;S277P;M441V;K497R;Y535H;I543L;S558L;T608S |  | PA<br>A/Netherlands/219/<br>2003(H7N7) |
| A/CATTLE/TEXAS<br>/81518/2024(H5N1)<br>PB1 | PB1 | T59S;E75D;M171V;V179I;N375S;R430E;A587P | N375S | PB1<br>A/Mallard/Astrakha<br>n/263/1982(H14N5) |
| A/CATTLE/TEXAS<br>/81518/2024(H5N1)<br>PB2 | PB2 | T58A;V109I;V139I;E362G;K389R;D441N;V478I;V495I;M631L;V649I;M676A | V495I;M676A | PB2<br>A/Duck/Guangdong<br>/E1/2012(H10N8) |
| A/CATTLE/TEXAS<br>/31156/2024(H5N1)<br>HA | HA | K3N;G16S;N110S;L120M;L131Q;T139P;T156A;Q185R;V194I;A201E;T211I;V226A;N252D;E284G;M285V;I298V;K492E;I526V;V538A;I547M;V548M | N110S;L131Q;<br>T139P;V226A | HA<br>A/Sichuan/26221/20<br>14(H5N6) |
| A/CATTLE/TEXAS<br>/31156/2024(H5N1)<br>M2 | M1 | N82S;N85S;N87T;T140A;F144L;M165I;V166A;A200V;A227T;K230R;M248L |  | M1<br>A/Duck/Guangdong<br>/E1/2012(H10N8) |
| A/CATTLE/TEXAS<br>/31156/2024(H5N1)<br>M2 | M2 | R12K;K18N;I51V;R61G;D88N |  | M2<br>A/Mallard/Astrakha<br>n/263/1982(H14N5) |
| A/CATTLE/TEXAS<br>/31156/2024(H5N1)<br>NA | NA | I8T;V17I;I20V;H44Y;A46P;V67I;N71S;T76A;K78Q;A81T;V99I;H100Y;H155Y;T188I;M258I;L269M;E287D;T289M;V321I;G336S;V338M;S339P;P340S;N366S;G382E;A395E;I418M;S434N;D460S |  | NA<br>A/Goose/Guangdon<br>g/1/1996(H5N1) |
| A/CATTLE/TEXAS<br>/31156/2024(H5N1)<br>NEP | NS1 | S7L;R21Q;S87P;C116S;D139N;A223E;V226I | A223E;V226I | NS1<br>A/Duck/Guangdong<br>/E1/2012(H10N8) |
| A/CATTLE/TEXAS<br>/31156/2024(H5N1)<br>NEP | NS2 | R61K;E67G | E67G | NS2<br>A/Duck/Guangdong<br>/E1/2012(H10N8) |
| A/CATTLE/TEXAS<br>/31156/2024(H5N1)<br>NP | NP | Y52H;S482N |  | NP<br>A/Chicken/BCFAV8<br>//2014(H5N2) |
| A/CATTLE/TEXAS<br>/31156/2024(H5N1)<br>PA | PA | I61M;T85A;K113R;L219I;S277P;M441V;K497R;Y535H;I543L;S558L;T608S |  | PA<br>A/Netherlands/219/<br>2003(H7N7) |
| A/CATTLE/TEXAS<br>/31156/2024(H5N1)<br>PB1 | PB1 | T59S;E75D;M171V;V179I;N375S;R430E;A587P | N375S | PB1<br>A/Mallard/Astrakha<br>n/263/1982(H14N5) |
| A/CATTLE/TEXAS<br>/31156/2024(H5N1)<br>PB2 | PB2 | T58A;V109I;V139I;E362G;K389R;D441N;V478I;V495I;M631L;V649I;M676A | V495I;M676A | PB2<br>A/Duck/Guangdong<br>/E1/2012(H10N8) |
| A/GRACKLE/TEX<br>AS/2/2024(H5N1)<br>HA | HA | K3N;G16S;N110S;L120M;L131Q;T139P;T156A;Q185R;V194I;A201E;T211I;V226A;N252D;E<br>284G;M285V;I298V;K492E;I526V;V538A;I547M;V548M | N110S;L131Q;<br>T139P;V226A | HA<br>A/Sichuan/26221/20<br>14(H5N6) |
| A/GRACKLE/TEX<br>AS/2/2024(H5N1)<br>M2 | M1 | N82S;N85S;N87T;T140A;F144L;M165I;V166A;A200V;A227T;K230R;M248L |  | M1<br>A/Duck/Guangdong<br>/E1/2012(H10N8) |
| A/GRACKLE/TEX<br>AS/2/2024(H5N1)<br>M2 | M2 | R12K;K18N;I51V;R61G;D88N |  | M2<br>A/Mallard/Astrakha<br>n/263/1982(H14N5) |
| A/GRACKLE/TEX<br>AS/2/2024(H5N1)<br>NA | NA | I8T;V17I;I20V;H44Y;A46P;V67I;N71S;T76A;K78Q;A81T;V99I;H100Y;H155Y;T188I;M258I;L<br>269M;E287D;T289M;V321I;G336S;V338M;S339P;P340S;N366S;G382E;A395E;I418M;S434N<br>;D460S |  | NA<br>A/Goose/Guangdon<br>g/1/1996(H5N1) |
| A/GRACKLE/TEX<br>AS/2/2024(H5N1)<br>NEP | NS1 | S7L;R21Q;S87P;C116S;D139N;A223E;V226I | A223E;V226I | NS1<br>A/Duck/Guangdong<br>/E1/2012(H10N8) |
| A/GRACKLE/TEX<br>AS/2/2024(H5N1)<br>NEP | NS2 | R61K;E67G | E67G | NS2<br>A/Duck/Guangdong<br>/E1/2012(H10N8) |
| A/GRACKLE/TEX<br>AS/2/2024(H5N1)<br>NP | NP | Y52H;S482N |  | NP<br>A/Chicken/BCFAV8<br>//2014(H5N2) |
| A/GRACKLE/TEX<br>AS/2/2024(H5N1)<br>PA | PA | I61M;T85A;K113R;L219I;S277P;M441V;K497R;Y535H;I543L;S558L;T608S |  | PA<br>A/Netherlands/219/<br>2003(H7N7) |
| A/GRACKLE/TEX<br>AS/2/2024(H5N1)<br>PB1 | PB1 | T59S;E75D;M171V;V179I;N375S;R430E;A587P | N375S | PB1<br>A/Mallard/Astrakha<br>n/263/1982(H14N5) |
| A/GRACKLE/TEX<br>AS/2/2024(H5N1)<br>PB2 | PB2 | T58A;V109I;V139I;E362G;K389R;D441N;V478I;V495I;M631L;V649I;M676A | V495I;M676A | PB2<br>A/Duck/Guangdong<br>/E1/2012(H10N8) |

